# Genetic evidence that high BMI in childhood has a protective effect on intermediate diabetes traits, including measures of insulin sensitivity and secretion

**DOI:** 10.1101/2023.02.03.23285420

**Authors:** Gareth Hawkes, Robin N Beaumont, Jessica Tyrrell, Grace M Power, Andrew Wood, Markku Laakso, Lilian Fernandes Silva, Michael Boehnke, Xianyong Yin, Tom G Richardson, George Davey Smith, Timothy M Frayling

## Abstract

Determining how high body-mass index (BMI) at different time points influences the risk of developing type two diabetes (T2D), and affects insulin secretion and insulin sensitivity, is critical. By estimating childhood BMI in 441,761 individuals in the UK Biobank, we identified which genetic variants had larger effects on adulthood BMI than on childhood BMI, and vice-versa. All genome-wide significant genetic variants were then used to separate the independent genetic effects of high childhood BMI from high adulthood BMI on the risk of T2D and insulin related phenotypes using Mendelian randomisation and studies of T2D, and oral and intravenous measures of insulin secretion and sensitivity. We found that a 1.s.d. (= 1.97kg/m^2^) higher childhood BMI, corrected for the independent genetic liability to adulthood BMI, was associated with a protective effect for seven measures of insulin sensitivity and secretion, including an increased insulin sensitivity index (β = 0.15 [0.067, 0.225], p = 2.79×10^−4^), and reduced fasting glucose (β = -0.053 [-0.089, -0.017], p = 4.31×10^−3^). There was however little to no evidence of a direct protective effect on T2D (OR = 0.94 [0.85 - 1.04], p = 0.228), independently of genetic liability to adulthood BMI. Our results thus cumulatively provide evidence of the protective effect of higher childhood BMI on insulin secretion and sensitivity, which are crucial intermediate diabetes traits. However, we stress that our results should not currently lead to any change in public health or clinical practice, given the uncertainty in biological pathway of these effects, and the limitations of this type of study.

**Research in Context:**

- High BMI in adulthood is associated with higher risk of type two diabetes, coupled with lower insulin sensitivity and secretion.
- Richardson et al [2020] used genetics to show that high BMI in childhood does not appear to increase the risk of type diabetes independently from its effect on adult BMI.
- We asked: does high childhood BMI affect insulin related traits such as fasting glucose and insulin sensitivity, independently of adulthood BMI?
- We used genetics to show that high childhood BMI has a protective effect on seven insulin sensitivity and secretion traits, including fasting glucose and measures of insulin sensitivity and secretion, independently of adulthood BMI.
- Our work has the potential to turn conventional understanding on its head – high BMI in childhood improves insulin sensitivity (when adjusting for knock on effects to high adult BMI) and opens up important questions about plasticity in childhood and compensatory mechanisms.

## Introduction

The increasing prevalence of obesity in childhood is assumed to lead to increased prevalence of type 2 diabetes in adult life [1]. Previous observational studies have shown that changing from a relatively thin child to overweight or obese adult provides additional risk to type 2 diabetes, compared to current adult BMI [2]. However, observational studies are subject to confounding which is less likely to impact genetic studies [3]. For example, an un-measured factor, such as smoking status, could act to confound the association between observed BMI and diabetes status, but can not affect the genetic variants that an individual carries.

A previous study that used genetics to understand the causality of higher BMI at different time points on T2D found that the relationship between childhood BMI and T2D was mediated through adulthood BMI [4]. That study used genetic variants with stronger effects on adulthood BMI than childhood BMI, and vice-versa, to separately test the effect of high BMI in childhood and the effects of high BMI in adulthood. However, that study was limited to analysis of type 2 diabetes as a binary disease trait, and did not investigate potential intermediate mechanisms such as those involving insulin secretion and sensitivity, and their genetic analysis was limited to lower-powered categorical BMI phenotypes. To understand more about the relationship between higher childhood BMI and type 2 diabetes, we generated a continuous measure of childhood BMI in the UK Biobank, validated using the 1958 National Childhood Development Study, which can be directly compared with continuous adulthood BMI, resulting in a more powerful genetic approach. We then tested a wide range of intermediate diabetes risk factors.

Using a combination of both previously identified and novel genetic instruments for childhood and adulthood BMI that resulted from our continuous phenotypes, we assessed the causal relationships between BMI at different life stages and diabetes outcomes: T2D, fasting insulin, fasting glucose and several measures of insulin secretion and sensitivity based on oral and intravenous tests, using Multivariable Mendelian Randomisation [5].

## Methods

### Study population

We analysed 441,761 individuals of calculated European descent within the UK Biobank (UKB) based on genetic principal component analysis, as previously described [6], with imputed genome-wide genetic variants, and both a baseline (adulthood) BMI measure (UKB Field 21001) and a self-recall variable related to body size at age 10 (UKB Field 1687).

### 1958 National Child Development Study

The 1958 National Child Development Study (1958NCDS) is a longitudinal assessment of 17,415 individuals who were born within a single week in March 1958. Beginning at the week of birth, mothers and their children were repeatedly assessed at irregular intervals, with a comprehensive set of measurements and assessments taken regarding many aspects of their lives (for more details see [7]). We analysed a subset of 5,847 individuals who had genome-wide array based genotyping and imputation, and were inferred to be of European descent again using genetic principal component analysis. Of these 5,847 individuals, 4,838 had measures of BMI at age 7, 4,704 at age 11, 4,298 at age 16, 5013 at age 23 and 5,774 at age 44.

### Measures of BMI

Individuals in the UKB were asked were asked whether they felt they were “thinner”, “the same size as” or “plumper” than their peers at age 10 (UKB Field 1687). From this categorical variable, we generated a continuous simulation of childhood BMI in the UKB based on summary statistics for BMI at age 11 from the 1958 National Child Development Study - see Supp Methods for full details.

Briefly, the self-recall variable for each participant’s body size at age 10 was used as an anchor to which we assigned an individual a BMI at age 10, after sub-sampling from a distribution which was an approximation of age 11 BMI in the 1958 NCDS. Next, using the UK Biobank Study, a genome-wide association study was performed for both adulthood and childhood BMI, from which we generated two polygenic scores. Adulthood BMI was taken directly from UKB Field 21001.

### Genetic Variants associated with BMI

We used the software *regenie* [8] to assess the association between each of 65,433,624 imputed genetic variants and BMI at each of the two timepoints independently for n=441,762 individuals. We then excluded genetic variants which were not single nucleotide polymorphisms, and those which did not have INFO*>* 0.8 and minor allele frequency (MAF) of 0.01*<*MAF*<*0.99. *regenie* performs association tests with a linear mixed model approach, which takes account of the degree of genetic relationship between each pair of individuals. BMI at both time points was rank inverse normalised and residualised at run time: covariates adjusted for were sex, age at baseline, genotyping chip and UKB assessment centre. As such, effect sizes are in standard deviation units.

Based on the results of these GWAS, we used Plink’s (v1.9) [9] clumping procedure to select independent and genome-level associated genetic variants. We used the following criteria to define an independent genetic association: *r*^2^ ≤ 0.001 (correlation between independent signals), dist≥ 250*kb* (distance between independent signals), *p* ≤ 5 × 10^−8^, 0.01 ≤ MAF ≤ 0.99, using an unrelated QC’d HapMap3 reference panel.

### Validation of Genetic Scores

The independent genetic variants for childhood and adulthood BMI derived here were individually assessed against phenotypes available in 1958NCDS. A genetic risk score (GRS), for both adulthood and age 10 BMI was calculated within the 1958NCDS cohort using the variants identified from the relevant GWAS, and assessed against derived BMI for each individual at ages 44 and 11. To make a direct comparison between the predictive ability of the two GRS’s against the two phenotypes, we calculated the receiver-operator-curves (ROCs) for the accuracy in predicting one of the two phenotypes being greater than 1s.d. from the standardized mean of the phenotype. We additionally calculate the percentage of variance explained by the continuous genetic score against the phenotype of interest.

At a genome-wide level, we calculated the genetic correlation between both adulthood and childhood BMI against both the most recent EGG-Consortium meta-analysis of childhood BMI [10] and adulthood BMI measured in [11]. We used the software R-package *LDSC* to calculate genetic correlation [12]. Finally, we also calculated the total variance explained by the instruments for adulthood and childhood BMI separately under the following formula: 2 × *β*^2^ × *MAF* × (1 − *MAF*), where MAF is the minor allele frequency.

### Mendelian Randomization

We used Mendelian Randomisation (MR) to assess whether there is a causal link [13] between our BMI exposures and T2D and insulin-related outcomes. In an MR study, the effect sizes of independent genetic variants that are strongly associated with each exposure are regressed against the effect sizes of the same variants with the disease/outcome from a secondary non-overlapping cohort’s GWAS (two sample MR). Comparison at a genetic level bypasses some observational confounders, as genetic variant genotypes are determined at zygote formation [13]. As such, an association found using MR provides stronger evidence of causality than that from observational data, albeit with some remaining confounding because of GWAS methodology.

We calculated the MR causal effect estimates using an inverse-variance weighted model, where each variant-exposure versus variant-outcome relationship is weighted by the inverse of the variance of the variant-outcome relationship. A sensitivity analysis was also performed in a lower power MR-Egger framework, which is more robust to pleiotropy (an association between the variant and outcome which does not pass through the exposure, which is a violation of the MR assumptions). Additionally, we performed a sensitivity analysis with Steiger filtering applied [14]. This approach excludes genetic variants that have larger effects on the outcomes (insulin secretion and sensitivity measures) than the exposures (childhood or adult BMI). We calculated effects using the formula used to compare instrument strength in the previous section. We also calculated an F-statistic for each of our analyses, as a measure of the quality of the variants as a genetic proxy for the observed exposure (typically F*>* 10 is classified as sufficient).

Where the variant-outcome relationship was not available in the outcome GWAS, a variant proxy was chosen based on a high degree of correlation (*r*^2^ *>* 0.8) between the index variant and its proxy, and a maximum distance between the index and proxy variant of 250kbp. Effect sizes between the variant and outcome, and variant-exposure were then either orientated to the matching alleles, or matched based upon the reported allele frequencies.

Multivariable mendelian randomisation (MVMR) is performed by conditioning the exposure-outcome relationship for each genetic variant upon that of another exposure’s genetic effect size (for example, conditioning the exposure-outcome relationship of childhood BMI upon that of adulthood BMI [15]). As such, the adjusted primary exposure-outcome relationship is independent of the genetic effects associated with the secondary exposure.

### Measures of T2D and insulin-related traits

Genetic variant effect sizes for T2D were drawn from [16], which was a meta-analysis of 71,124 cases and 824,006 controls of European ancestry and FinnGen (Freeze Six) [17] for “Type 2 diabetes, strict (exclude DM1)”, which included 37,002 cases and 215,160 controls. The MR results for each T2D outcome GWAS were then meta-analysed.

Effect sizes relating to fasting glucose (FG) and fasting insulin (FI) were drawn from [18], where FG was measured in 151,188 individuals and FI in 105,056 individuals.

We also analysed seven measures of insulin sensitivity and response during and after an oral glucose tolerance test in a meta-analysis of results from [19] (n = 26,037 participants without diabetes) and the METSIM study (n = 8,520). Specifically, we looked at the association of our BMI measures with the area under the insulin curve (AUC), the ratio of AUCs for insulin and glucose (AUC ratio), an insulin sensitivity index (ISI), insulin after 30 minutes (Ins 30), insulin after 30 minutes adjusted for BMI (Ins 30 adj BMI), incremental insulin relative to fasting insulin after 30 minutes (Incremental Ins 30) and corrected insulin response (CIR) - see [19] for specific definitions.

Meta-analyses were performed, where applicable, using the ‘*metafor*’ R package, based on the assumption of a fixed effect between the exposure and outcome across studies.

## Results

In our analysis of 441,762 adult individuals (of ages from 40 to 75 years) we identified 306 (Supp Table 1) and 1127 (Supp Table 2) independent genetic variants (*p <* 5 × 10^−8^, 250kb distance) associated with continuous measures of childhood and adulthood BMI respectively, in comparison to [4], where 295 and 557 independent signals were reported respectively for categorical measures of the same outcomes. The exact parameters derived to describe the continuous measure of childhood BMI are given in the Supplementary Results. The variance explained by the genetic variants were 11.3% and 4.03% for adulthood and childhood BMI, compared to 2.78% and 1.96% in Richardson et. al, demonstrating that our genetic instruments have been strengthened by using continuous variables.

Using these variants we generated polygenic scores for childhood and adulthood BMI, and validated them in the 1958 dataset and independent data from the EGG and GIANT consortia. The adulthood BMI GRS was a better predictor of standardised adulthood BMI (age 44) being greater than one standard deivation from the mean in the 1958 dataset (OR = 1.55 [1.43-1.67] p = 1.29×10^−16^, var = 4.80%), than of standardised childhood BMI (age 11) being more than one standard deviation from the mean in the same dataset (OR = 1.32 [1.20 - 1.44], p = 1.85×10^−9^, var = 0.995%), and explained more of the variance (‘var’) in the respective continuous traits - Supp Fig 4. The childhood BMI GRS was a better predictor of childhood BMI (OR = 1.32 [1.21 - 1.44], p = 1.32×10^−16^, var = 1.62%) than of adulthood (OR = 1.16 [1.08 - 1.25], p =3.73×10^−5^, var = 0.635%).

The genetic correlation between adulthood BMI measured in this UK Biobank study, and childhood BMI measured directly by the EGG consortium (N = 35,668, ages 2 to 10 years), was 0.644 [0.582 - 0.705] (p = 3.22×10^−95^), which was less than that between our measure of childhood BMI and the EGG consortium, at 0.937 [0.864 - 1.01] (p = 1.29×10^−145^). The genetic correlation between adulthood BMI analysed in [11] by the GIANT consortium and adulthood BMI measured here was 0.943 [0.925 - 0.960], p = 1.31×10^−210^, versus the genetic correlation with childhood BMI measured here at 0.645 [0.591 - 0.680], p = 1.28×10^−173^.

## Childhood BMI

Using MR we showed that higher BMI in childhood was associated with protective effects on diabetes related traits after adjustment for the independent effects of higher adulthood BMI. The results are as follows:

### Mendelian Randomisation showed that higher Childhood BMI, corrected for genetic liability to adulthood BMI, has protective effects on measures on insulin secretion and sensitivity traits

In a multivariable model which takes account of the independent genetic effects of adulthood BMI, we found that a 1s.d. (=1.97kg/m^2^) higher childhood BMI was associated with a protective effect on a range of insulin secretion and sensitivity traits: lower area under the insulin curve (AUC), ratio of insulin and glucose curves (AUC Ratio), insulin after 30 minutes adjusted for BMI (Ins 30 BMI adj), insulin change after 30 minutes (Incr Ins 30), and higher insulin sensitivity index (ISI) (Figure 2 & Supp Table 3) - see methods for precise trait definitions. Higher childhood BMI was also weakly associated with lower FG (β = -0.0528 [-0.0893, -0.0163], p = 4.21×10^−3^). There was no evidence of an effect on FI (β = -0.0109 [-0.0564, 0.0346], p = 0.638).

**Fig 1.**
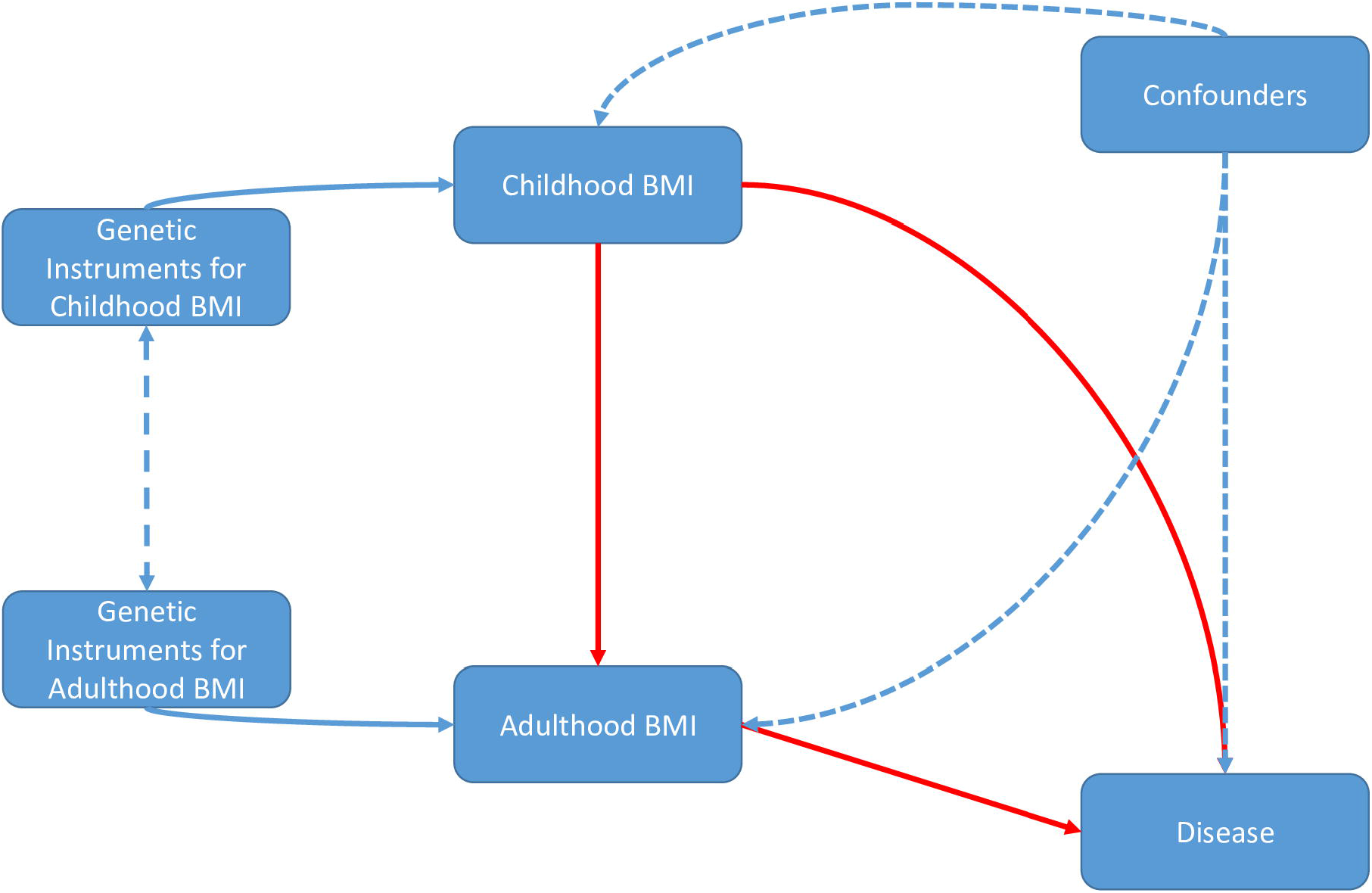
Directed acyclic graph illustrating assumed causal relationship between childhood and adulthood. Solid red lines denote causality, unidirectional blue dashes represent non-causal effects, and double-ended blue dashed lines denote a covariance structure.

**Fig 2.**
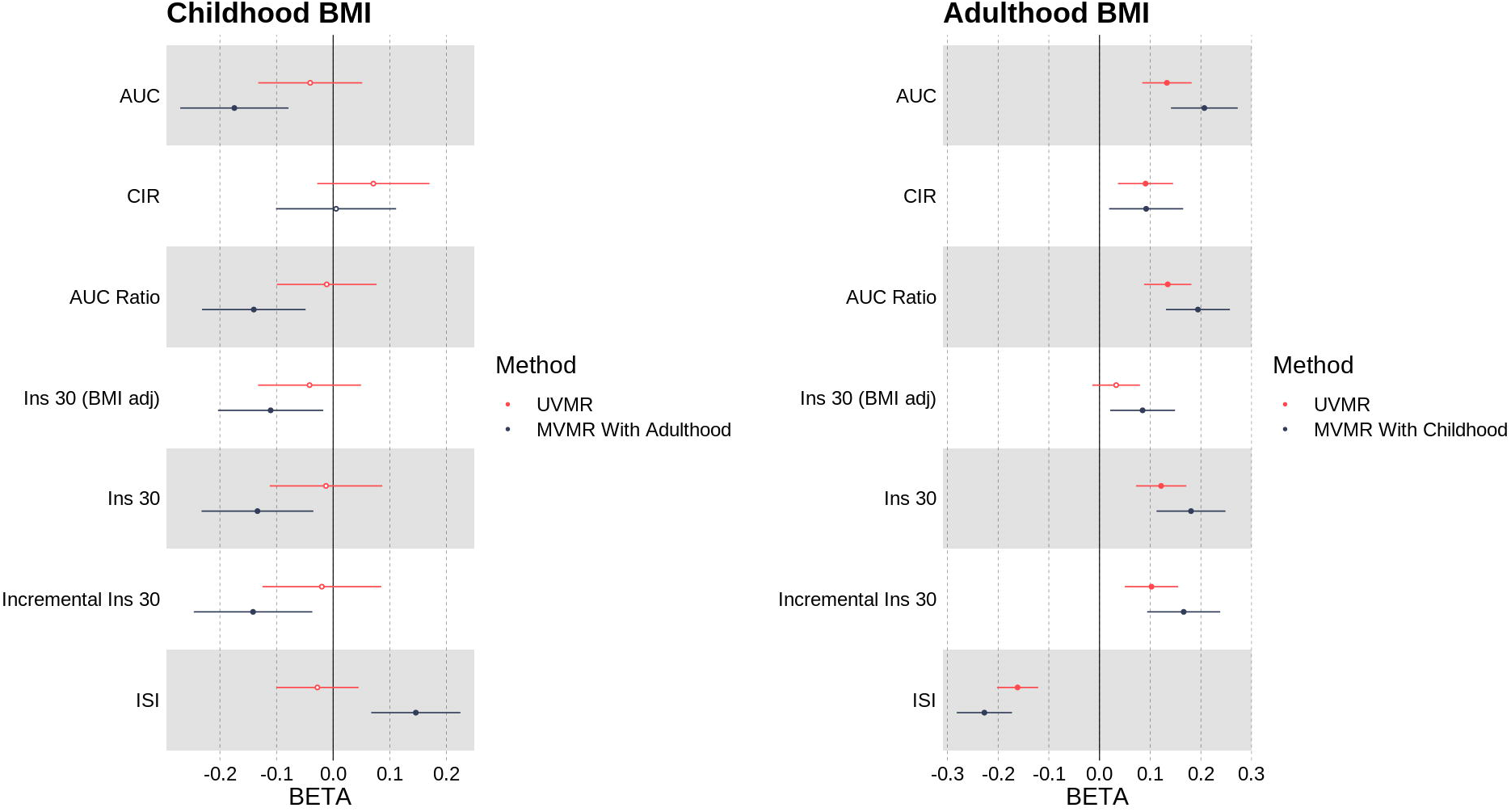
Univariable (MR) and multivariable (MVMR) meta-analysis results for childhood and adulthood BMI versus oral glucose tolerance test traits: area under the insulin curve (AUC), ratio of insulin and glucose curves (AUC Ratio), insulin after 30 minutes adjusted for BMI (Ins 30 BMI adj), insulin change after 30 minutes (Incr Ins 30), and higher insulin sensitivity index (ISI)

These results are shown, including the comparison to univariable models with no adjustment for adulthood BMI, in Figs 2 and 3, and Supp Table 3.

**Fig 3.**
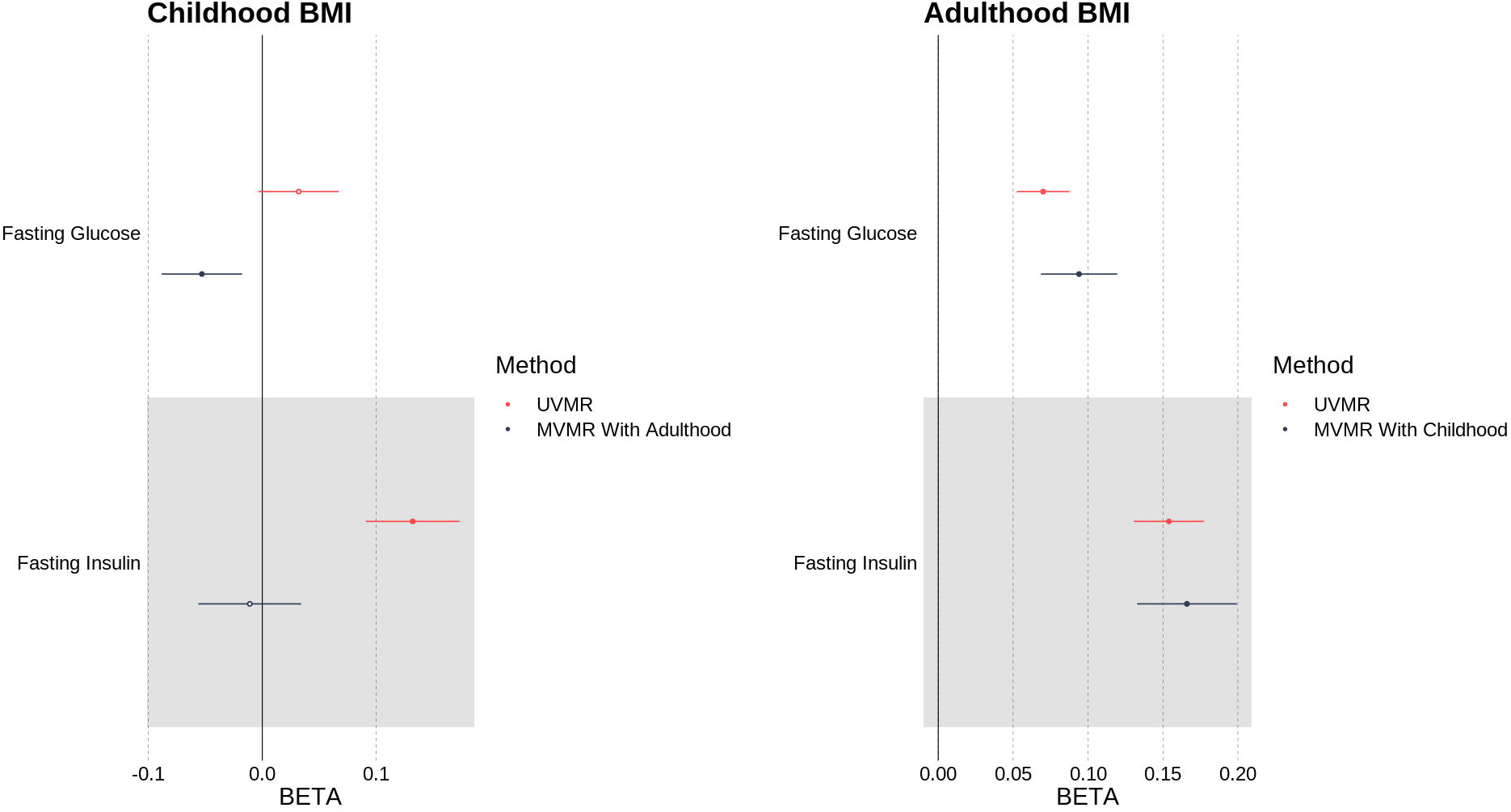
Univariable (MR) and multivariable (MVMR) results of testing the association between childhood and adulthood BMI against fasting insulin (FI) and fasting glucose (FG).

**Fig 4.**
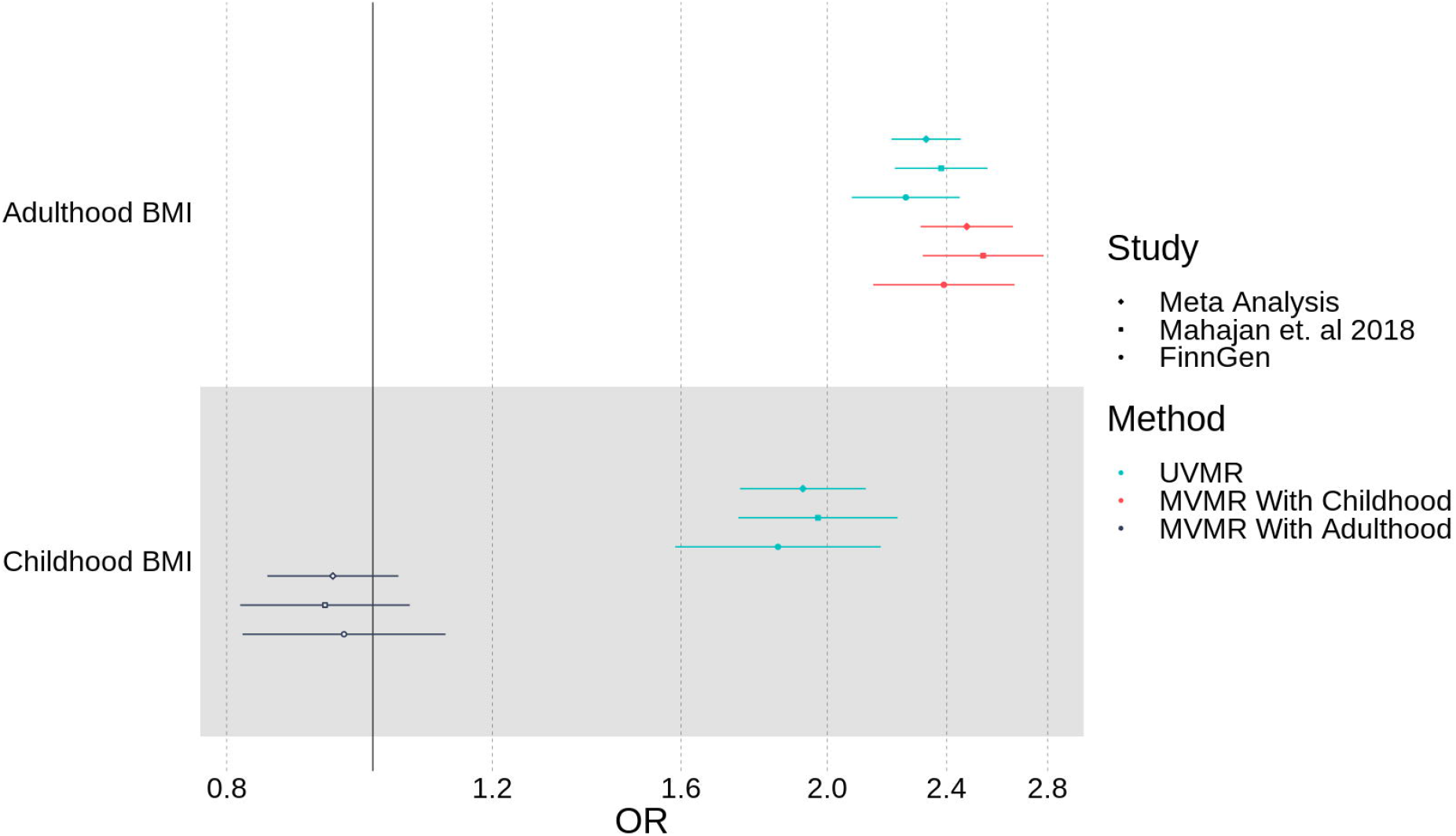
Univariable (MR) and multivariable (MVMR) results of association between childhood and adulthood BMI against T2D as measured in Mahajan et. al 2018 and FinnGen, with meta-analysis.

### Mendelian Randomization showed that higher Childhood BMI, corrected for the genetic liability to adulthood BMI, was not protective for T2D

In a multivariable model which corrects for the independent genetic liability to adulthood BMI, higher childhood BMI was not associated (OR = 0.941 [0.851, 1.04], p = 0.228) with the risk of T2D, consistent across two meta-analysed studies [13, 16]. These results are shown in comparison to the univariable model with no adjustment for adulthood BMI, in Fig 4, and Supp Table 3.

## Adulthood BMI

Using MVMR, we showed that higher BMI in adulthood leads to a higher risk of T2D, independently of the genetic effects childhood BMI. We observed consistent effects on insulin and glycaemic traits intermediate to T2D, with MVMR showing that higher adult BMI leads to lower insulin sensitivity and higher insulin secretion. The link between higher genetically derived adult BMI and higher insulin secretion in people without T2D is likely a response to lower insulin sensitivity.

### Mendelian Randomization showed that higher Adulthood BMI was associated with a damaging effect on insulin and glycaemic traits

In a multivariable model which corrects for the independent genetic liability to childhood BMI, a 1s.d. (=4.77kg/m^2^) higher adulthood BMI was associated with higher levels of fasting glucose (FG) (β = 0.0941 [0.0683, 0.120], p = 7.50×10^−13^) and fasting insulin (FI) (β = 0.166 [0.134, 0.199], p = 3.19×10^−10^). An increase in adulthood BMI in a multivariable model also showed evidence of a damaging effect on the remaining six insulin traits (Supp Table 3). These results, in comparison to the univariable model with no adjustment for childhood BMI, are shown in Figs 2 and 3, and Supp Table 3.

### Mendelian Randomization showed that higher Adulthood BMI was associated with an increased risk of T2D

In a multivariable model which corrects for the independent genetic liability to childhood BMI, a 1s.d. (=4.77kg/m^2^) higher adulthood BMI was associated with an increased risk of T2D (OR = 2.47 [2.31, 2.65], p = 1.23×10^−142^). These results, in comparison to the univariable model with no adjustment for childhood BMI, are shown in Figs 4, and Supp Table 3.

### Sensitivity Analyses

MR-Egger intercept analyses for each of the 57 MVMR models identified 3 statistical associations with p<0.01 in the following exposure-outcome relationships: fasting insulin from the MAGIC consortium (β = 1.40×10^−3^, p = 9.44×10^−4^), and T2D from both the FinnGen and Mahajan et. al studies (β = (4.00×10^−3^, 4.00×10^−3^) and p = (5.00×10^−3^, 1.00×10^−3^) respectively) – see Supp Table 4.

A Steiger filtered analysis resulted in consistently wider confidence intervals as compared to those without filtering – see Supp Table 5, but the two sets of effect sizes were highly correlated (r = 0.947) and were directionally consistent in 101/106 analyses (MR and MVMR).

## Discussion

We have used genetics and Mendelian randomization to assess the independent causal relationships between BMI recalled from childhood and adulthood against the risk of developing T2D, as well as their effects on insulin-related traits using fasting, oral glucose tolerance test and intravenous glucose tolerance tests. Our measure of childhood BMI provided a more powerful genetic measure than previous work, based upon a combination of self-recall categories and known summary statistics from the 1958 National Child Development Study for measured childhood BMI.

We found using genetics that higher BMI in childhood, once separated from higher BMI in adulthood, was protective for measures of both insulin secretion and sensitivity, including fasting glucose and an insulin sensitivity index. We note that associations with lower insulin secretion when not corrected for insulin sensitivity are consistent with a protective effect - because, in people without diabetes, a higher insulin sensitivity results in reduced need for insulin secretion. One possible explanation is that higher adiposity in childhood stimulates differentiation of cells important for insulin sensitivity and secretion - such as adipocytes and beta-cells. There is evidence that people with higher BMIs but without diabetes have more beta cells [20], and evidence that more adipocytes are present in people without diabetes compared to those with diabetes but of the same BMI [21]. However, the evidence that higher childhood BMI leads to a protective effect on measures of insulin secretion and sensitivity was not reflected in a conclusive association with protection from T2D. This difference may be due to differences in power between measures of continuous traits and binary disease traits, although the T2D sample sizes are larger than those for the intermediate traits. Additionally, this effect is relative to other people who may have changed BMI between childhood and adulthood - for example, it may be that change in BMI is the true risk factor, resulting in higher BMI in childhood appearing less damaging than a lower BMI in childhood, the latter of which would lead to greater relative increases in adulthood. It is also possible that the genetic variants with stronger associations with childhood BMI result primarily in higher muscle or non-fat mass components to growth. Associations with higher muscle mass could result in higher insulin sensitivity and help explain our findings. There is, however, recent work suggesting that the childhood genetic variants used in [4], and that overlap strongly with the genetic variants we used, were good measures of fat mass in children at age 9 to 18 [22]. Crucially, they found that the genetic variants were more strongly associated with adiposity than lean mass in childhood to the beginning of adulthood. The study showed that the variants derived from the UKB childhood BMI recall variable are more strongly associated with adiposity than lean mass as measured by DEXA imaging measures at ages 9,13,15, and 25. The data showed convincingly that the higher childhood BMI genetic score leads to higher adiposity consistently at several time points in childhood after the adiposity rebound at age 4-5 years and that these effects are stronger with fat mass than lean mass, although both are present. The associations were also consistent with trajectories of BMI in childhood. More precisely, the childhood BMI genetic instrument was associated with consistently stronger effects on the imaging-based measures than the adulthood genetic instrument at age 9, 13,15 and 18 years, with a 1 SD higher childhood BMI genetic risk score associated with ∼8% higher fat mass compared to 1-2% higher lean mass. The adulthood genetic instrument had a stronger effect on fat mass than the childhood instrument by age 25 years. It is possible that the likely insulin sensitizing effects of the 1-2% higher lean mass at several times points aged 9-18 offset the insulin resistance effects of the 8% higher fat mass. However, we think this is unlikely because we know that a higher fat mass leads to slightly higher lean mass due to the load bearing effects of the extra weight, as seen with the *FTO* variant [23]. Importantly the adult BMI genetic instrument is associated with a proportionally similar increase in lean mass for each 1 SD increase in fat mass, and we know this genetic instrument is associated with lower insulin sensitivity. Whilst we cannot rule it out, we therefore think it unlikely that non-fat mass effects and different trajectories of growth are influencing our results. Because of these uncertainties, we stress that our work should not lead to any change in clinical practice during childhood, early life or adulthood, and more work is needed to identify the biological mechanisms that could be driving these associations.

Adulthood BMI was found to have a consistently risk increasing/damaging effect on all traits studied, regardless of whether the independent genetic liability to childhood BMI was corrected for, acting as a positive control.

There are a few notable limitations to this study. First, our measure for childhood BMI is derived from a categorical measure which was recalled many decades after the truth. To attempt to overcome this limitation, we performed validation in both the 1958 Birth Cohort, and against external GWAS of BMI measured in childhood, where we found that our genetic measures are more strongly predictive of the chronologically correct phenotypes.

We were unable to independently certify that the genetic variants we have used as a proxy of childhood BMI were associated with early-life adiposity, as opposed to (for example) growth and lean mass. We also acknowledge the limitations related to an MR study, where fully satisfying the three fundamental assumptions is rarely achieved. For example, an MR analysis assumes that the genetic variant does not affect the outcome other than via the exposure: this is unlikely to be consistently the case when considering genetic variants which increase the odds of having T2D and BMI, if the variant (for example) raised insulin sensitivity independently. There was also some evidence of pleiotropy for 3 of our analyses using the MR-Egger test.

In summary, our data provides initial evidence that higher fat mass in childhood leads to relative protective effects – improvements in insulin sensitivity and reduced need for insulin secretion – in adulthood. A potential explanation is the beneficial effects of exposure to the metabolic challenges of higher adiposity in early, more plastic, stages of life compared to the likely damaging effects of large increases in adiposity between childhood and adulthood.

## Data Availability

Data cannot be shared publicly because of data availability and data return policies of the UK Biobank. Data are available from the UK Biobank for researchers who meet the criteria for access to datasets to UK Biobank (http://www.ukbiobank.ac.uk).

## Acknowledgments

We want to acknowledge the participants and investigators of the FinnGen, METSIM, 1958NCDS and UK Biobank studies. This manuscript is part of the Stratification of Obesity Phenotypes to Optimize Future Obesity Therapy (SOPHIA) project. SOPHIA has received funding from the Innovative Medicines Initiative 2 Joint Undertaking under grant agreement No. 875534. This Joint Undertaking support from the European Union’s Horizon 2020 research and innovation program and EFPIA and T1D Exchange, JDRF, and Obesity Action Coalition www.imisophia.eu. GH has received funding from the Innovative Medicines Initiative 2 Joint Undertaking under grant agreement No 875534. JT is supported by an Academy of Medical Sciences (AMS) Springboard award, which is supported by the AMS, the Wellcome Trust, GCRF, the Government Department of Business, Energy and Industrial strategy, the British Heart Foundation and Diabetes UK [SBF004\1079]. TGR and GDS work within the MRC Integrative Epidemiology Unit at the University of Bristol supported by the Medical Research Council (MC UU 00011/1). The research utilised data from the UK Biobank resource carried out under UK Biobank application number 9072. UK Biobank protocols were approved by the National Research Ethics Service Committee. The authors would like to acknowledge the Exeter Sequencing service in carrying out the RNA-Sequencing. The equipment utilised is funded by the Wellcome Trust Institutional Strategic Support Fund (WT097835MF), Wellcome Trust Multi User Equipment Award (WT101650MA) and BBSRC LOLA award (BB/K003240/1). The authors would like to acknowledge the use of the University of Exeter High-Performance Computing (HPC) facility in carrying out this work. We acknowledge use of high-performance computing funded by an MRC Clinical Research Infrastructure award (MRC Grant: MR/M008924/1). TMF is supported by MRC awards MR/WO14548/1 and MR/T002239/1. MB, XY, and ML are supported by NIH grant DK062370.

## Disclaimer

This communication reflects the author’s views: neither IMI nor the European Union, EFPIA, or any Associated Partners are responsible for any use that may be made of the information contained therein.

## Notes

### Competing Interest Statement

The authors have declared no competing interest.

### Author Declarations

This research has been conducted using the UK Biobank Resource. This work was carried out under UK Biobank project number 9072. Ethics approval for the UK Biobank study was obtained from the North West Centre for Research Ethics Committee (11/NW/0382) [38]. Written informed consent was obtained from all participants. The study was performed in accordance with the Declaration of Helsinki.

